# Greater Effect of Polygenic Risk Score for Alzheimer’s Disease Among Younger Cases who are Apolipoprotein E-ε4 Carriers

**DOI:** 10.1101/2020.04.06.20052332

**Authors:** Brian Fulton-Howard, Alison M. Goate, Robert P. Adelson, Jeremy Koppel, Marc L. Gordon, Alzheimer’s Disease Genetics Consortium, Nir Barzilai, Gil Atzmon, Peter Davies, Yun Freudenberg-Hua

## Abstract

To evaluate how age and Apolipoprotein E-ε4 (*APOE4)* status interact with *APOE-* independent polygenic risk score (PRS_non-APOE_), we estimated PRS_non-APOE_ in superagers (age ≥ 90 years, N=346), 89- controls (age 60-89, N=2,930) and Alzheimer’s Disease (AD) cases (N=1,760). Employing superagers, we see a nearly five times greater odds ratio (OR) for AD comparing the top PRS_non-APOE_ decile to the lowest decile (OR=4.82, P=2.5×10^-6^), which is twice the OR as using 89- controls (OR=2.38, P=4.6×10^-9^). Thus PRS_non-APOE_ is correlated with age, which in turn is associated with *APOE*. Further exploring these relationships, we find that PRS_non-APOE_ modifies age-at-onset among *APOE4* carriers, but not among non-carriers. More specifically, PRS_non-APOE_ in the top decile predicts an age-at-onset five years earlier compared to the lowest decile (70.1 vs 75.0 years; *t*-test P=2.4×10^-5^) among *APOE4* carriers. This disproportionally large PRS_non-APOE_ among younger *APOE4*-positive cases is reflected in a significant statistical interaction between *APOE4* status and age-at-onset (β=-0.02, P=4.8×10^-3^) as a predictor of PRS_non-APOE_. Thus, the known AD risk variants are particularly detrimental in young *APOE4* carriers.

**Disclosure Statement:** AMG has consulted for Eisai, Biogen, Pfizer, AbbVie, Cognition Therapeutics and GSK. She also served on the SAB at Denali Therapeutics from 2015-2018. YFH co-owns stock and stock options of Regeneron Pharmaceuticals. All other authors have no interests to declare.

## 1. Introduction

Alzheimer’s disease (AD) has an estimated heritability of 59-79% (Gatz et al., 2006) and a decades long pre-clinical stage, during which AD-specific pathological changes develop without clinical signs of cognitive decline (Bateman et al., 2012; Jack et al., 2010). Low-cost non-invasive testing would ideally identify at-risk individuals in the pre-clinical stage to enable preventive intervention. To this end, genetic risk factors have been proposed as useful predictors. *APOE* is a major risk gene for AD with dose-dependent risk effects. Compared to non-carriers of the *APOE-*ε4 allele (*APOE4)*, heterozygous carriers of *APOE4* have a three- to fourfold and homozygous carriers up to a 14-fold increased odds of developing AD (Farrer et al., 1997; Genin et al., 2011), whereas the *APOE-*ε2 allele (*APOE2*) is protective (odds ratio, OR=0.6) (Bertram et al., 2008; Bickeböller et al., 1997; Corder et al., 1994; Genin et al., 2011; Kunkle et al., 2019a). The effects of *APOE* variants have been found to be age-dependent (Rasmussen et al., 2018). In addition to the *APOE* gene, genome-wide association studies (GWAS) and meta-analysis of GWAS have identified 39 risk loci in subjects with European ancestry (Jansen et al., 2019; Kunkle et al., 2019b; Lambert et al., 2013b; Marioni et al., 2018). Desikan et al. constructed a polygenic hazard score (PHS) by combining AD-associated single nucleotide polymorphisms (SNPs) from GWAS from the International Genomics of Alzheimer’s Project (IGAP) and disease incidence estimates from the United States population (Desikan et al., 2017). This PHS was reported to be predictive for age-at-onset (AAO) and useful to identify individuals at greatest risk for developing AD at a given age. However, the interactions between *APOE*-independent genetic risks, age and *APOE* have not been evaluated. Similar to PHS, individual genetic risk can be aggregated into a polygenic risk score (PRS) by summing the number of risk alleles, each weighted by its effect size derived from independent GWAS summary statistics (Euesden et al., 2015; Khera et al., 2018; Purcell et al., 2009). Both PRS and PHS have been applied to AD and showed similar results (Leonenko et al., 2019) although a conflicting study (Fan et al., 2020) reported that PHS is superior for predicting age at onset and other age-related phenotypes, particularly when stratified by sex. PRS, rather than PHS, is broadly used and may be considered the current state of the art for genetic risk prediction. In addition, because PRS quantifies genetic risk in individuals independently of age, PRS can also be used to explore the interactions between genetics and other causes of disease, including age. PRS has been reported to be useful to identify individuals at high risk for common diseases such as coronary artery disease (Khera et al., 2018). For AD, the predictive power of PRS derived from the GWAS meta-analysis (Lambert et al., 2013a) reached an area under the curve (AUC) of 0.78 in clinically-defined AD when genetic risk factors were combined with demographic factors, and 0.84 in pathologically-confirmed AD (Escott-Price et al., 2017; Escott-Price et al., 2015).

The predictive performance of PRS was shown to depend on the prevalence of the disease in a specific population (Gibson et al., 2019), which is meaningful for AD, given that prevalence of AD increases exponentially with age (Mayeux and Stern, 2012). Furthermore, it is important to note that *APOE* is not only a major constituent for predicting AD (Desikan et al., 2017; Escott-Price et al., 2015), but is also strongly associated with age itself (Deelen et al., 2019; Sebastiani et al., 2019) as well as AAO of AD (Olarte et al., 2006). Therefore, it is important to investigate whether and how *APOE*-independent PRS (PRS_non-APOE_) depends on age and *APOE*, both for the interpretation of AD PRS and to understand how genetic risk is distributed in case-control AD genetics studies.

To address this question, we stratified healthy controls into superagers aged 90 years or older without dementia (90+; range 90-109 years) and subjects aged 89 or younger (89-; range 60-89 years). To understand the role of age with regard to PRS_non-APOE_, we compared these two control cohorts to AD cases with a broad range of AAO (range 60-99 years). We further investigated whether PRS_non-APOE_ is correlated with AAO in cases and age of controls and whether the *APOE4* risk allele would modify such a correlation.

## 2. Methods

### 2.1 Participants

To ensure that there is no overlap with the samples included in the 2013 IGAP meta-analysis (Lambert et al., 2013b), we selected 13 independent cohorts consisting of a total of 1,733 cases and 3,121 controls from the Alzheimer’s Disease Genetics Consortium (ADGC) (**Supplementary Methods**). Additionally, we included an independent sample of 182 subjects with Ashkenazi ancestry from the Litwin Zucker Alzheimer’s Research Center at the Feinstein Institutes for Medical Research (LZ). The sample ascertainment and assessment methods were described previously (Adelson et al., 2019; Freudenberg-Hua et al., 2016; Koppel et al., 2018). A visual overview of data preparation and analysis workflow is in **Supplementary Figure 1**.

Age was defined using ADGC criteria. For ADGC cases, age is defined as AAO as reported by the study (n=1,574). If AAO is not reported, we estimated AAO by subtracting ten years from age at death (AAD, n=102) and for those without AAD, age at last examination (AAE) is used (n=57). AAO estimation has been used in previous studies using the same cohorts (Desikan et al., 2017; Jun et al., 2010; Naj et al., 2014). To ensure that estimated AAO does not introduce biases, we further performed sensitivity analysis by removing individuals with estimated AAO from the dataset. For the ADGC controls, AAE is used (n=2,995) and AAD is used for a small proportion of controls (n=126). For the LZ cohort, age is reported as age at enrollment for cases (n=27), and as AAE for controls (89-: n=35, 90+: n=120). The total sample utilized in this study includes 1,760 AD cases (60-99 years), 2,930 89- controls, and 346 90+ controls. Sample and genotype data quality control procedures and principal component analysis are described in **Supplementary Methods**. The demographics of the sample set following merging and QC are shown in **Supplementary Table 1**. The distribution of *APOE* genotypes among cases and controls is described in **Supplementary Table 2**.

### 2.2 Polygenic Risk Score Estimation

A PRS is a sum of genetic variants, weighted by variant effect sizes for a given trait. The effect sizes are estimated from independent GWAS summary statistics. PRS_non-APOE_ is calculated at various P-value thresholds (P_T_) based on GWAS summary statistics (Lambert et al., 2013c) excluding the *APOE* region (**Supplementary Methods**). We separately optimized PRS_non-APOE_ using two control sets – ADGC controls without age restriction and 90+ controls (**Supplementary Figure 2**) – which resulted in identical optimal P_T_ for the most predictive PRS at P_T_ =1×10^-5^. Details of PRS calculation are in **Supplementary Methods**.

### 2.3 Statistical Analysis

Statistical analysis was performed using the R software version 3.6.3 (R Core Team, 2019). Linear or logistic regression models included covariates for sex and the top ten principal components (PCs), as well as the variables of interest. All plots and statistical tests (except for those in **Figure 1a**) used *Z*-standardized PRS. To facilitate comparison with previous findings on polygenic risks and survival, we plotted a Kaplan-Meier survival curve with the primary event being AAO of AD and we calculated hazard ratio using the R packages “survival” and “survminer”.

**Figure 1.**
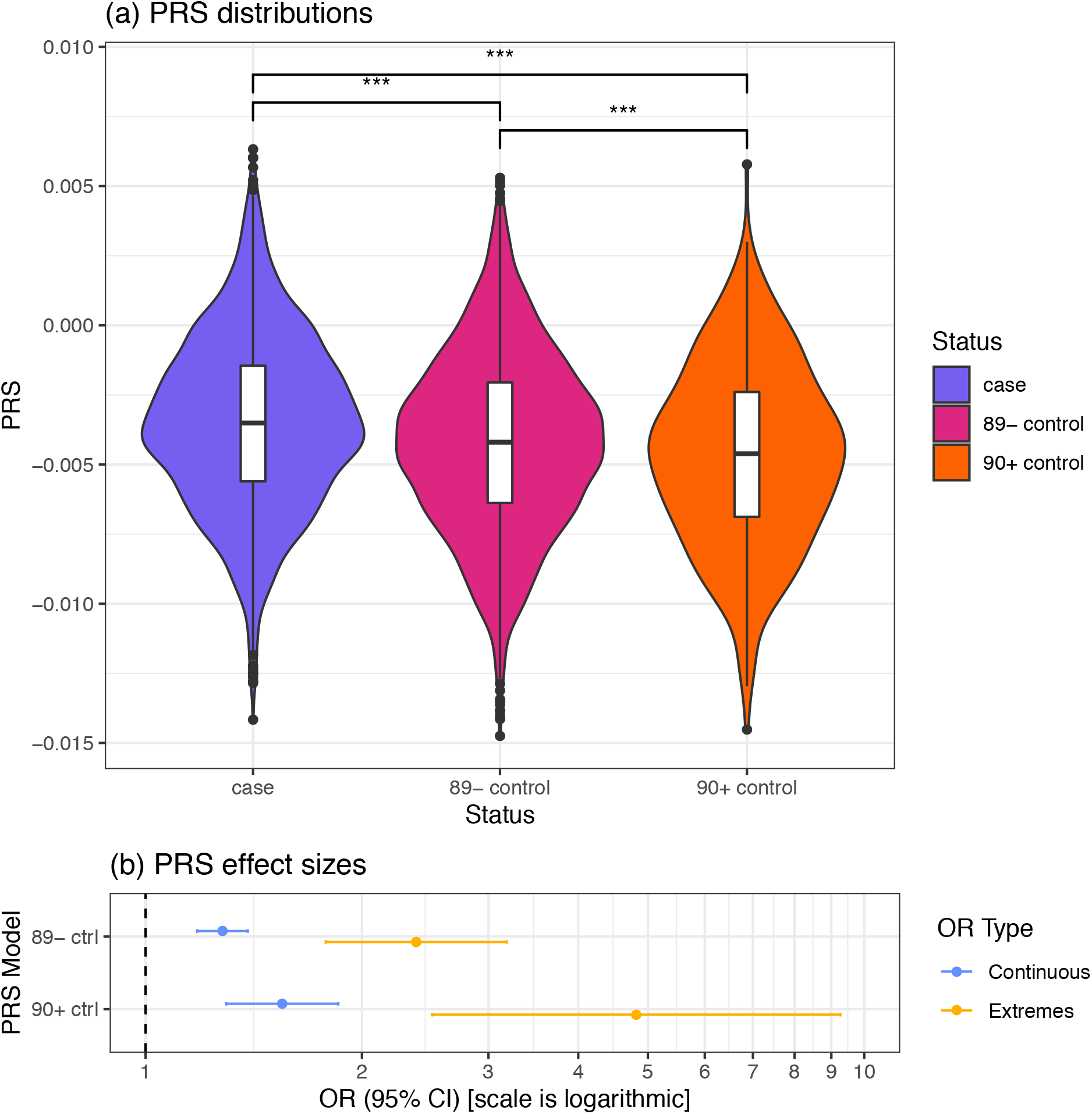
PRS_non-APOE_ Distribution and Effect Sizes in the AD Case and Control Groups (Residualized). **(a)**, Violin plots comparing PRS_non-APOE_ in AD cases (case), controls age 60-89 years (89-), and superager controls age ≥ 90 years (90+). Pairwise comparison Bonferroni P-values: *** highly significant (P < 0.001). **(b)**, PRS effects on AD risk. Continuous OR (blue): odds ratio of having AD for a one SD increase in PRS_non-APOE_. Extremes OR (orange): odds ratio of having AD with PRS_non-APOE_ in the top decile compared to having PRS_non-APOE_ in the lowest decile. Abbreviations: 90+ ctrl, controls ≥ 90 years old were used in analysis; 89- ctrl, controls age 60-89; AD, Alzheimer’s disease; CI, confidence interval; OR, odds ratio; PRS_non-APOE_, *APOE*-independent polygenic risk score.

To compare PRS distributions across cases, 89- controls, and 90+ controls, we performed a one-way ANOVA and pairwise Student’s *t*-tests. In order to ensure that observed differences were not due to ethnicity or sex, we tested the residuals of a linear model with sex and ten PCs as predictors and PRS as the response variable.

We evaluated the association between PRS and AD status in two datasets: 1) cases and 89- controls; and 2) cases and superager (90+) controls. To determine the difference of odds ratio (OR) for AD between the extremes of PRS, we selected the bottom and top deciles of the PRS distribution and performed logistic regressions between PRS decile and AD status (extremes model). To determine the effect of a one standard deviation (SD) increase in PRS, we performed logistic regressions with PRS – z-standardized across all samples – as the predictor and AD status as the response (continuous model). The effect size of this continuous model is useful because it does not depend on the extremeness of PRS quantization (e.g. top and bottom 10% vs. 5%).

It has been previously shown that polygenic risk predicts AAO of AD. In this study we further investigate how age and *APOE4* status modify the effects of PRS. Therefore, we modeled the association between age, *APOE* genotype (i.e., 2/2, 2/3, 3/4, or 4/4, dummied with *APOE3*/*3* as the contrast variable), and PRS using linear regression, with covariates for confounding variables as before. To determine if these effects are independent of AD status, we performed an additional linear regression controlling for AD status. We also tested for collinearity by calculating the variance inflation factor (VIF) with the “car” package.

To test the interaction between age and *APOE* genotype on PRS, we stratified by AD case-control status, categorized samples as *APOE4* carriers and non-carriers, and then tested for interactions. To improve interpretability by reducing the number of interactions tested, we collapsed the *APOE* genotypes into binary categories – an *APOE4* carrier was defined as having *APOE3*/*4* or *4*/*4* (excluding *2*/*4*), and a non-carrier as *APOE2*/*2*, *2*/*3* or *3*/*3*. We first performed linear regressions separately in cases and controls, with *APOE4* carrier status and age as predictors and PRS as the response. We then performed a second linear regression stratified by AD status, adding the interaction between *APOE4* carrier status and age as a predictor.

Finally, to test for the effect of these interactions on case-control status, we performed logistic regressions for the relationship between AD case-control status, *APOE4* carrier status, age, and PRS, with and without interactions and confirmed independence of predictors with VIF.

## 3. Results

### 3.1 Superagers have the lowest PRS_non-APOE_

PRS_non-APOE_ is different between cases, 89- controls, and 90+ controls after residualizing for sex and ancestry PCs (ANOVA P=1×10^-19^) (**Figure 1a**). AD subjects have significantly higher PRS_non-APOE_ compared to both 89- controls (Bonferroni adj. post-hoc *t*-test, P=5.5×10^-4^) and 90+ controls (P=3.6×10^-20^). The 89- controls have significantly higher PRS_non-APOE_ than 90+ controls (P=5.1×10^-14^). This difference shows the dependence of PRS_non-APOE_ on age among healthy controls. To further evaluate this age dependence, we assessed the effect of PRS_non-APOE_ on AD risk by comparing cases with 89- controls and then with 90+ controls (**Figure 1b**). We first calculated “extremes” ORs between subjects in the bottom and top deciles of PRS_non-APOE_. Comparing 90+ controls and AD cases, subjects having PRS_non-APOE_ in the top decile are nearly five times more likely to have AD than subjects with PRS_non-APOE_ in the lowest decile (OR=4.82, CI: 2.50-9.26, P=2.5×10^-6^). This effect size is double the odds ratio using 89- controls (OR=2.38, CI: 1.78-3.18, P=4.6×10^-9^). This difference remains present with the alternative approach of “continuous” ORs measuring the effects of a one SD increase in PRS_non-APOE_, albeit less pronounced (90+ controls: OR=1.55, CI: 1.29-1.85, P=1.8×10^-6^; 89-controls: OR=1.28, CI: 1.18-1.39, P=2.1×10^-9^). The higher effect sizes of PRS_non-APOE_ observed when comparing with 90+ controls are consistent with the effect sizes seen for the *APOE4* alleles and genotypes (**Supplementary Table 3**).

### 3.2 The negative association between age and PRS_non-APOE_ interacts with *APOE* genotype in cases

The particularly low PRS_non-APOE_ observed in superagers led us to elucidate the relationship between PRS and age. Biologically, PRS can affect age of inclusion or onset in AD studies by modifying both AD risk and AAO. Here, we investigated age as a predictor for PRS to explore how non-APOE genetic predictors change with age in case-control studies, and how *APOE* genotype affects those changes. Age is negatively correlated with PRS_non-APOE_ among all subjects (**Supplementary Table 4**). In a linear regression including sex, *APOE* and ten PCs, age is a significant predictor of PRS_non-_ APOE (P=2.7×10^-5^, β=-0.007). This effect remains significant when accounting for case-control status (P=1.6×10^-4^, β =-0.006) (**Supplementary Table 4**). No *APOE* genotype is directly associated with PRS_non-APOE_.

We next stratified the subjects into *APOE4* carriers and non-carriers to investigate whether *APOE4* carrier status modifies the correlation between age and PRS_non-APOE_. We find that this correlation depends on both case-control status and *APOE4* status (**Figure 2a**). Interestingly, the negative correlation between age and PRS_non-APOE_ depends on *APOE4* carrier status in cases, but not in controls (**Figure 2b-c, Supplementary Table 5)**. More specifically, in AD cases, AAO is correlated with PRS_non-APOE_ (P=0.029, β=-0.007) and *APOE4* carrier status is not associated with PRS_non-APOE_ (P=0.58). However, when the interaction between AAO and *APOE* (P=4.8×10^-3^, β=-0.02) is accounted for, *APOE4* status becomes a significant predictor of PRS_non-APOE_ (P=6.1×10^-3^, β=1.29), whereas age is no longer a significant predictor (P=0.72). Thus, the inverse correlation between AAO and PRS_non-APOE_ among cases is primarily driven by the relationship between PRS_non-APOE_ on AAO among *APOE4* carriers (**Figure 2b**). Consequently, among AD cases who are *APOE4* carriers, those with PRS_non-APOE_ in the top decile have an AAO five years earlier than those in the bottom decile (70.1 vs. 75.0 years; 95% CI of difference in means: −7.2 – −2.7; P=2.4×10^-5^). This difference in AAO remains significant after removing *APOE4/4* homozygous cases (71.1 vs. 76.7 years; P=2.1×10^-5^). A sensitivity analysis removing cases with estimated AAO further shows that these interaction results are robust (Supplementary Table 6).

**Figure 2.**
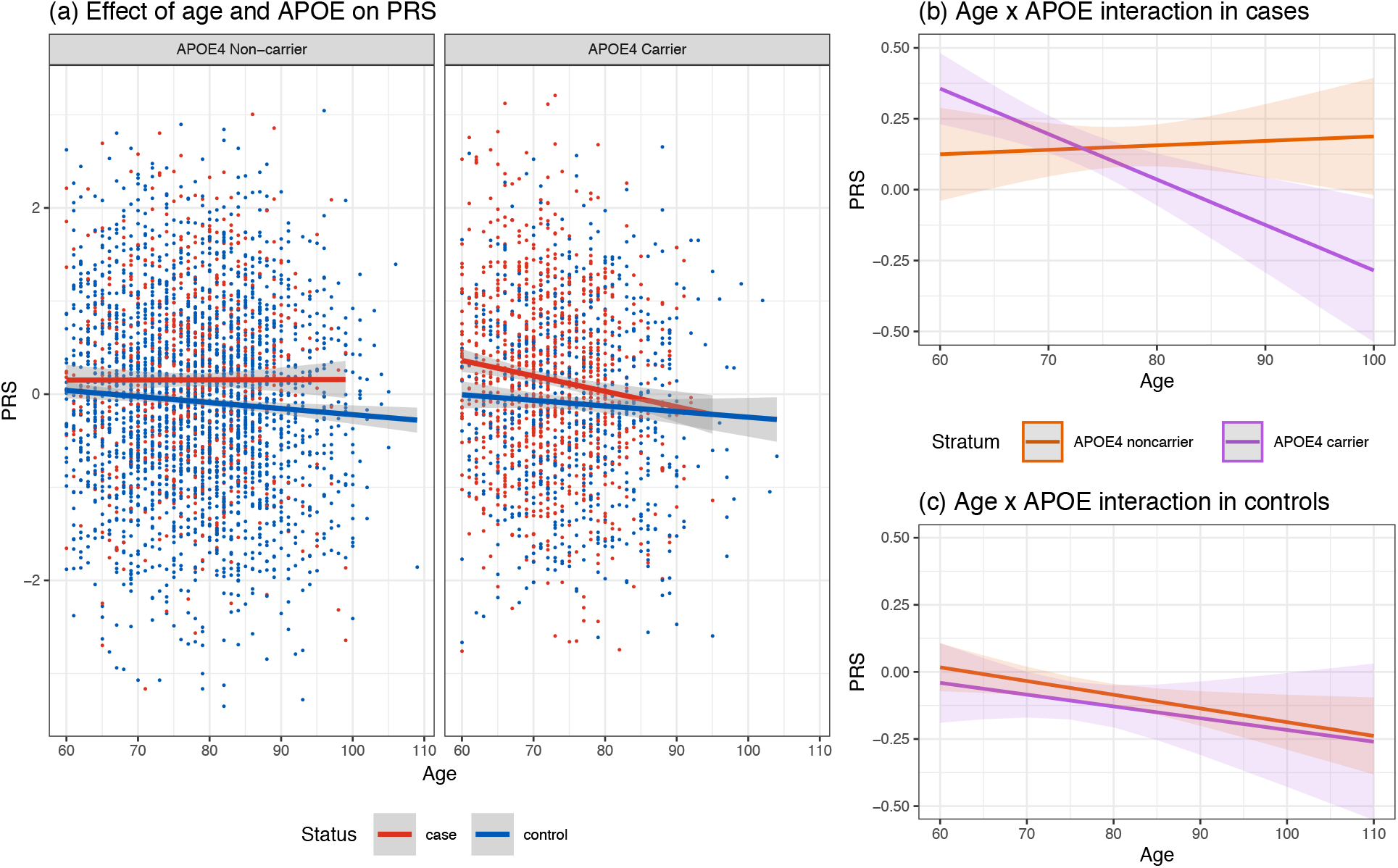
Regression Plots Visualizing Interactions Between PRS, *APOE4*, and Age, in Cases and Controls. (**a**) Scatter plots and regression models showing the dependence of PRS_non-APOE_ on age in AD cases (red) and controls (blue). In AD cases age is defined as age-at-onset. The shaded areas show the 95% confidence intervals. PRS_non-APOE_ in cases and controls over age is shown separately among *APOE4* non-carriers and carriers. (**b**) Age x *APOE* interaction in AD cases. (**c**) Age x *APOE* interaction in controls, which is negatively associated with PRS for both *APOE4* carriers (purple) and non-carriers (orange). Abbreviations: AD, Alzheimer’s disease; *APOE4*, Apolipoprotein E-ε4 allele; PRS_non-APOE_, *APOE*-independent polygenic risk score.

In contrast, the negative correlation between age and PRS_non-APOE_ (P=0.013, β=-0.005) does not depend on *APOE4* carrier status among controls (**Figure 2c, Supplementary Table 5**), and *APOE* is not associated with PRS_non-APOE_ (P=0.27). Accounting for interactions between age and *APOE4* status, the negative association between age and PRS_non-APOE_ remains significant (P=0.021, β=-0.005) and *APOE4* status non-significant (P=0.78). The interaction between *APOE4* and age is not significant (P=0.88). It is worthwhile to note that the main effect sizes while accounting for interactions do not have any intuitive interpretation due to the extrapolation of the age variable (Aschard, 2016).

### 3.3 Interactions between age, PRS_non-APOE_, and *APOE* on AD status

The interactions between AAO and *APOE4* status led us to evaluate the effect of the interactions between *APOE4* status, age and PRS_non-APOE_ on AD status using logistic regression (**Supplementary Table 7**). Without modeling for interactions, age (P=2.4×10^-6^, β=-0.02), *APOE4* carrier status (P=8.6×10^-86^, β=1.31) and PRS_non-APOE_ (P=2.7×10^-12^, β=0.23) are all predictors of AD status. When all interactions between these three predictors are included in the model, the main effects of age (P=0.60) and PRS_non-APOE_ (P=0.41) are no longer significant, indicating that the effects of age and PRS_non-APOE_ on AD status are dependent on *APOE*. *APOE4* carrier status remains a significant predictor (P=8.2×10^-15^, β=4.79). Similarly, the main effects of the interactions between age and *APOE* (P=1.3×10^-8^, β=-0.05), between PRS_non-APOE_ and *APOE* (P=0.035, β=1.31), and the three-way interaction between age, PRS and *APOE* (P=0.035, β=- 0.02) are all significant predictors for AD status.

The Receiver Operator Characteristics (ROC) are similar to previous reports when *APOE* SNPs are included (**Supplementary Figure 3**) and the Kaplan-Meier survival curve for PRS_non-APOE_ stratified by *APOE4* carriers status is similar to that previously reported (**Supplementary Figure 4**) (Desikan et al., 2017).

## 4. Discussion

We show that *APOE*-independent polygenic risk has a disproportionally large effect in younger *APOE4* carriers. Among *APOE4* carriers, the AAO is on average five years earlier for cases with PRS_non-APOE_ in the top decile compared to those with PRS_non-APOE_ in the lowest decile. In contrast, no correlation between PRS_non-APOE_ and AAO was observed among *APOE4* non-carriers. We further observe a negative correlation between age and PRS_non-APOE_ that is independent of *APOE4* carrier status among controls, consistent with the notion that *APOE*-independent genetic risk increases AD incidence throughout late life. Our findings indicate that younger *APOE4* carriers bear greater detrimental effect from currently known AD risk variants, as captured by PRS_non-APOE_, whereas superager controls have fewer AD risk variants independent from *APOE*. Even though the age effect per year is small, it can be considerable in a population of subjects with ages ranging from 60 to over 100 years. This pattern may either be inherent to genetically conferred risk for AD or reflect biases in current PRS estimates as a result of ascertainment of previous GWAS subjects.

Compared with healthy superagers, subjects having PRS_non-APOE_ in the top decile have nearly five times the odds of developing AD than those in the bottom decile. This large effect size may be caused by depletion of AD risk alleles among superagers, indicating that many typical controls will develop AD later in life. This is consistent with the long pre-clinical stage of 20 to 25 years during which a subject does not display dementia symptoms (Bateman et al., 2012). Our observation also corroborates the reported higher effect sizes of GWAS risk alleles when superagers are used as controls (Tesi et al., 2019). The lifetime risk of AD for healthy females and males at 75 years old is estimated to be 26.2% and 19.9%, respectively (Brookmeyer et al., 2018). As the majority of controls enrolled in AD genetic studies are in their 70s and 80s, a considerable proportion of them are expected to convert to AD at a later age unless they die from other causes. These cryptic preclinical AD cases reduce the power of case-control studies. The larger magnitude of effect sizes for both PRS_non-APOE_ and *APOE* when 90+ controls were deployed in our study highlights the importance of inclusion of cognitively healthy superagers in future studies. Increasing the number of superager controls in case-control studies for AD is expected to increase the power to detect additional risk variants as well as protective variants.

The greater PRS_non-APOE_ among younger *APOE4* positive cases, as reflected in the statistical interaction between AAO and *APOE4* carrier status, suggests a greater role for the currently known *APOE*-independent genetic risk in younger *APOE4* carriers. Although PRS_non-APOE_ alone is not useful to clinically predict individual risk for developing AD – perhaps unlike PHS/PRS including APOE (Desikan et al., 2017; Escott-Price et al., 2015) – the interactions we observed have implications for the design of future genetic association studies. In particular, enrollment of *APOE4*-negative cases irrespective of age may lead to the identification of novel variants, given the absent correlation of current PRS_non-APOE_ with AAO in this group. Novel non-genetic protective factors may best be found among healthy elderly *APOE4* carriers with high PRS_non-APOE_ based on known GWAS risk loci, whereas novel risk factors may best be found among AD patients who have both low PRS_non-APOE_ and no *APOE4* alleles.

Recently another study, investigating subjects that do not overlap with our study and using different parameters, reported a greater effect of *APOE* in younger participants (Bellou et al., 2020). Our stratified interaction analysis indicating a larger effect of PRS_non-APOE_ in younger *APOE4* carriers leads us to a novel conclusion about *APOE*-independent genetic risk and provides a logical explanation for their observation: among all *APOE4* cases, those with a younger onset carry the largest PRS_non-APOE_.

Our study has several limitations, some of which are inherently shared by many large-scale AD genetic studies. In order to reach sufficient power, genetic studies of complex diseases require very large sample size which is often achieved by pooling subjects from many studies that were initially independently designed. This approach introduces heterogeneity in diagnostic criteria, recorded demographics, and comorbidities (Jun et al., 2010; Naj et al., 2011). Such heterogeneity typically introduces noise and reduces statistical power for detecting true signals. There are now collaborative efforts by the National Institute on Aging to harmonize genetic, epidemiologic and clinical data for AD. Notwithstanding the importance of clinical details, genetic studies based solely on self-report of parental AD in the UK Biobank have yielded results that are highly correlated with the previously known IGAP loci (Andrews et al., 2020; Marioni et al., 2018). This indicates that genetic studies can generate valid findings without considering or correcting for all clinical details, when large sample sizes are used. In our study, AAO for AD was generally provided by the participating cohorts of ADGC (Desikan et al., 2017; Jun et al., 2010). Our sensitivity analysis shows that removing subjects with an estimated AAO does not substantially affect our overall results. However, a larger study may be required to fully measure the interactions between PRS_non-APOE_ and the number of *APOE4* and *APOE2* alleles. Additionally, because of differences across ethnicities in PRS applicability (Martin et al., 2019) and *APOE* effects (Rajabli et al., 2018), studies in non-European populations are necessary to determine whether the same conclusions apply.

## 5. Conclusion

In summary, we show that current *APOE*-independent PRS has a particularly detrimental effect in younger *APOE4* risk allele carriers, predicting significantly earlier age at onset for AD. *APOE*-independent PRS is negatively correlated with dementia-free aging. Future larger studies are needed to evaluate whether interactions between PRS, *APOE* and additional demographic, clinical, and environmental risk factors, as reflected in health data and biomarkers, can improve AD risk prediction.

## Data Availability

All genetic data referred to in the manuscript will be made available in a suitable repository, and is available upon reasonable request made to the corresponding author following peer-reviewed publication of the manuscript.

## Acknowledgements

Funding: This work was supported by the National Institutes of Health, National Institute on Aging (NIH-NIA) (ADGC and grant numbers U01 AG032984 and RC2 AG036528).

Samples from the National Cell Repository for Alzheimer’s Disease (NCRAD), which receives government support under a cooperative agreement awarded by the NIH-NIA (grant number U24 AG21886), were used in this study. The NACC database is funded by the NIH-NIA (grant number U01 AG016976). Data for this study were prepared, archived, and distributed by the National Institute on Aging Alzheimer’s Disease Data Storage Site (NIAGADS) at the University of Pennsylvania (grant number U24-AG041689-01). In addition, this work was funded by the NIH (grant numbers U01AG052411, U01AG058635, K08AG054727, R01 AG618381, and R01 AG046949), and the Einstein Nathan Shock Center (grant number P30 AG038072). Additional support was provided by the Mildred and Frank Feinberg Family Foundation, the Advancing Women in Science and Medicine Foundation, and the JPB Foundation.

The authors would like to thank the contributors who collected samples used in this study during a period of 30 years, as well as patients and their families, whose help and participation made this work possible. A list of ADGC members and their affiliations are included in the **Appendix** within the **Supplementary Content** file.

## References

Adelson, R.P., Renton, A.E., Li, W, Barzilai, N., Atzmon, G., Goate, A.M., Davies, P., Freudenberg-Hua, Y., 2019. Empirical design of a variant quality control pipeline for whole genome sequencing data using replicate discordance. Sci Rep 9(1), 16156.

Andrews, S.J., Fulton-Howard, B., Goate, A., 2020. Interpretation of risk loci from genome-wide association studies of Alzheimer’s disease. Lancet Neurol 19(4), 326–335.

Aschard, H., 2016. A perspective on interaction effects in genetic association studies. Genet Epidemiol 40(8), 678–688.

Bateman, R.J., Xiong, C., Benzinger, T.L., Fagan, A.M., Goate, A., Fox, N.C., Marcus, D.S., Cairns, N.J., Xie, X., Blazey, T.M., Holtzman, D.M., Santacruz, A., Buckles, V., Oliver, A., Moulder, K., Aisen, P.S., Ghetti, B., Klunk, W.E., McDade, E., Martins, R.N., Masters, C.L., Mayeux, R., Ringman, J.M., Rossor, M.N., Schofield, P.R., Sperling, R.A., Salloway, S., Morris, J.C., Network, D.I.A., 2012. Clinical and biomarker changes in dominantly inherited Alzheimer’s disease. N Engl J Med 367(9), 795–804.

Bellou, E., Baker, E., Leonenko, G., Bracher-Smith, M., Daunt, P., Menzies, G., Williams, J., Escott-Price, V., Initiative, A.s.D.N., 2020. Age-dependent effect of APOE and polygenic component on Alzheimer’s disease. Neurobiol Aging 93, 69-77.

Bertram, L., Lange, C., Mullin, K., Parkinson, M., Hsiao, M., Hogan, M.F., Schjeide, B.M., Hooli, B., Divito, J., Ionita, I., Jiang, H., Laird, N., Moscarillo, T., Ohlsen, K.L., Elliott, K., Wang, X., Hu-Lince, D., Ryder, M., Murphy, A., Wagner, S.L., Blacker, D., Becker, K.D., Tanzi, R.E., 2008. Genome-wide association analysis reveals putative Alzheimer’s disease susceptibility loci in addition to APOE. Am J Hum Genet 83(5), 623–632.

Bickeböller, H., Campion, D., Brice, A., Amouyel, P., Hannequin, D., Didierjean, O., Penet, C., Martin, C., Pérez-Tur, J., Michon, A., Dubois, B., Ledoze, F., Thomas-Anterion, C., Pasquier, F., Puel, M., Demonet, J.F., Moreaud, O., Babron, M.C., Meulien, D., Guez, D., Chartier-Harlin, M.C., Frebourg, T., Agid, Y., Martinez, M., Clerget-Darpoux, F., 1997. Apolipoprotein E and Alzheimer disease: genotype-specific risks by age and sex. Am J Hum Genet 60(2), 439–446.

Brookmeyer, R., Abdalla, N., Kawas, C.H., Corrada, M.M., 2018. Forecasting the prevalence of preclinical and clinical Alzheimer’s disease in the United States. Alzheimers Dement 14(2), 121–129.

Corder, E.H., Saunders, A.M., Risch, N.J., Strittmatter, W.J., Schmechel, D.E., Gaskell, P.C., Rimmler, J.B., Locke, P.A., Conneally, P.M., Schmader, K.E., 1994. Protective effect of apolipoprotein E type 2 allele for late onset Alzheimer disease. Nat Genet 7(2), 180–184.

Deelen, J., Evans, D.S., Arking, D.E., Tesi, N., Nygaard, M., Liu, X., Wojczynski, M.K., Biggs, M.L., van der Spek, A., Atzmon, G., Ware, E.B., Sarnowski, C., Smith, A.V., Seppälä, I., Cordell, H.J., Dose, J., Amin, N., Arnold, A.M., Ayers, K.L., Barzilai, N., Becker, E.J., Beekman, M., Blanché, H., Christensen, K., Christiansen, L., Collerton, J.C., Cubaynes, S., Cummings, S.R., Davies, K., Debrabant, B., Deleuze, J.F., Duncan, R., Faul, J.D., Franceschi, C., Galan, P., Gudnason, V., Harris, T.B., Huisman, M., Hurme, M.A., Jagger, C., Jansen, I., Jylhä, M., Kähönen, M., Karasik, D., Kardia, S.L.R., Kingston, A., Kirkwood, T.B.L., Launer, L.J., Lehtimäki, T., Lieb, W., Lyytikäinen, L.P., Martin-Ruiz, C., Min, J., Nebel, A., Newman, A.B., Nie, C., Nohr, E.A., Orwoll, E.S., Perls, T.T., Province, M.A., Psaty, B.M., Raitakari, O.T., Reinders, M.J.T., Robine, J.M., Rotter, J.I., Sebastiani, P., Smith, J., Sørensen, T.I.A., Taylor, K.D., Uitterlinden, A.G., van der Flier, W., van der Lee, S.J., van Duijn, C.M., van Heemst, D., Vaupel, J.W., Weir, D., Ye, K., Zeng, Y., Zheng, W., Holstege, H., Kiel, D.P., Lunetta, K.L., Slagboom, P.E., Murabito, J.M., 2019. A meta-analysis of genome-wide association studies identifies multiple longevity genes. Nat Commun 10(1), 3669.

Desikan, R.S., Fan, C.C., Wang, Y., Schork, A.J., Cabral, H.J., Cupples, L.A., Thompson, W.K., Besser, L., Kukull, W.A., Holland, D., Chen, C.H., Brewer, J.B., Karow, D.S., Kauppi, K., Witoelar, A., Karch, C.M., Bonham, L.W., Yokoyama, J.S., Rosen, H.J., Miller, B.L., Dillon, W.P., Wilson, D.M., Hess, C.P., Pericak-Vance, M., Haines, J.L., Farrer, L.A., Mayeux, R., Hardy, J., Goate, A.M., Hyman, B.T., Schellenberg, G.D., McEvoy, L.K., Andreassen, O.A., Dale, A.M., 2017. Genetic assessment of age-associated Alzheimer disease risk: Development and validation of a polygenic hazard score. PLoS Med 14(3), e1002258.

Escott-Price, V., Myers, A.J., Huentelman, M., Hardy, J., 2017. Polygenic risk score analysis of pathologically confirmed Alzheimer disease. Ann Neurol 82(2), 311–314.

Escott-Price, V., Sims, R., Bannister, C., Harold, D., Vronskaya, M., Majounie, E., Badarinarayan, N., Morgan, K., Passmore, P., Holmes, C., Powell, J., Brayne, C., Gill, M., Mead, S., Goate, A., Cruchaga, C., Lambert, J.C., van Duijn, C., Maier, W., Ramirez, A., Holmans, P., Jones, L., Hardy, J., Seshadri, S., Schellenberg, G.D., Amouyel, P., Williams, J., GERAD/PERADES, consortia, I., 2015. Common polygenic variation enhances risk prediction for Alzheimer’s disease. Brain 138(Pt 12), 3673-3684.

Euesden, J., Lewis, C.M., O’Reilly, P.F., 2015. PRSice: Polygenic Risk Score software. Bioinformatics 31(9), 1466–1468.

Fan, C.C., Banks, S.J., Thompson, W.K., Chen, C.H., McEvoy, L.K., Tan, C.H., Kukull, W., Bennett, D.A., Farrer, L.A., Mayeux, R., Schellenberg, G.D., Andreassen, O.A., Desikan, R., Dale, A.M., 2020. Sex-dependent autosomal effects on clinical progression of Alzheimer’s disease. Brain 143(7), 2272–2280.

Farrer, L.A., Cupples, L.A., Haines, J.L., Hyman, B., Kukull, W.A., Mayeux, R., Myers, R.H., Pericak-Vance, M.A., Risch, N., van Duijn, C.M., 1997. Effects of age, sex, and ethnicity on the association between apolipoprotein E genotype and Alzheimer disease. A meta-analysis. APOE and Alzheimer Disease Meta Analysis Consortium. JAMA 278(16), 1349–1356.

Freudenberg-Hua, Y., Li, W., Abhyankar, A., Vacic, V., Cortes, V., Ben-Avraham, D., Koppel, J., Greenwald, B., Germer, S., Darnell, R.B., Barzilai, N., Freudenberg, J., Atzmon, G., Davies, P., T2D-GENES Consortium, 2016. Differential burden of rare protein truncating variants in Alzheimer’s disease patients compared to centenarians. Hum Mol Genet 25(14), 3096–3105.

Gatz, M., Reynolds, C.A., Fratiglioni, L., Johansson, B., Mortimer, J.A., Berg, S., Fiske, A., Pedersen, N.L., 2006. Role of genes and environments for explaining Alzheimer disease. Arch Gen Psychiatry 63(2), 168–174.

Genin, E., Hannequin, D., Wallon, D., Sleegers, K., Hiltunen, M., Combarros, O., Bullido, M.J., Engelborghs, S., De Deyn, P., Berr, C., Pasquier, F., Dubois, B., Tognoni, G., Fievet, N., Brouwers, N., Bettens, K., Arosio, B., Coto, E., Del Zompo, M., Mateo, I., Epelbaum, J., Frank-Garcia, A., Helisalmi, S., Porcellini, E., Pilotto, A., Forti, P., Ferri, R., Scarpini, E., Siciliano, G., Solfrizzi, V., Sorbi, S., Spalletta, G., Valdivieso, F., Vepsalainen, S., Alvarez, V., Bosco, P., Mancuso, M., Panza, F., Nacmias, B., Bossù, P., Hanon, O., Piccardi, P., Annoni, G., Seripa, D., Galimberti, D., Licastro, F., Soininen, H., Dartigues, J.F., Kamboh, M.I., Van Broeckhoven, C., Lambert, J.C., Amouyel, P., Campion, D., 2011. APOE and Alzheimer disease: a major gene with semi-dominant inheritance. Mol Psychiatry 16(9), 903–907.

Gibson, J., Russ, T.C., Clarke, T.K., Howard, D.M., Hillary, R.F., Evans, K.L., Walker, R.M., Bermingham, M.L., Morris, S.W., Campbell, A., Hayward, C., Murray, A.D., Porteous, D.J., Horvath, S., Lu, A.T., McIntosh, A.M., Whalley, H.C., Marioni, R.E., 2019. A meta-analysis of genome-wide association studies of epigenetic age acceleration. PLoS Genet 15(11), e1008104.

Jack, C.R., Knopman, D.S., Jagust, W.J., Shaw, L.M., Aisen, P.S., Weiner, M.W., Petersen, R.C., Trojanowski, J.Q., 2010. Hypothetical model of dynamic biomarkers of the Alzheimer’s pathological cascade. Lancet Neurol 9(1), 119–128.

Jansen, I.E., Savage, J.E., Watanabe, K., Bryois, J., Williams, D.M., Steinberg, S., Sealock, J., Karlsson, I.K., Hägg, S., Athanasiu, L., Voyle, N., Proitsi, P., Witoelar, A., Stringer, S., Aarsland, D., Almdahl, I.S., Andersen, F., Bergh, S., Bettella, F., Bjornsson, S., Brækhus, A., Bråthen, G., de Leeuw, C., Desikan, R.S., Djurovic, S., Dumitrescu, L., Fladby, T., Hohman, T.J., Jonsson, P.V., Kiddle, S.J., Rongve, A., Saltvedt, I., Sando, S.B., Selbæk, G., Shoai, M., Skene, N.G., Snaedal, J., Stordal, E., Ulstein, I.D., Wang, Y., White, L.R., Hardy, J., Hjerling-Leffler, J., Sullivan, P.F., van der Flier, W.M., Dobson, R., Davis, L.K., Stefansson, H., Stefansson, K., Pedersen, N.L., Ripke, S., Andreassen, O.A., Posthuma, D., 2019. Genome-wide meta-analysis identifies new loci and functional pathways influencing Alzheimer’s disease risk. Nat Genet 51(3), 404–413.

Jun, G., Naj, A.C., Beecham, G.W., Wang, L.S., Buros, J., Gallins, P.J., Buxbaum, J.D., Ertekin-Taner, N., Fallin, M.D., Friedland, R., Inzelberg, R., Kramer, P., Rogaeva, E., St George-Hyslop, P., Cantwell, L.B., Dombroski, B.A., Saykin, A.J., Reiman, E.M., Bennett, D.A., Morris, J.C., Lunetta, K.L., Martin, E.R., Montine, T.J., Goate, A.M., Blacker, D., Tsuang, D.W., Beekly, D., Cupples, L.A., Hakonarson, H., Kukull, W., Foroud, T.M., Haines, J., Mayeux, R., Farrer, L.A., Pericak-Vance, M.A., Schellenberg, G.D., Consortium, A.s.D.G., 2010. Meta-analysis confirms CR1, CLU, and PICALM as alzheimer disease risk loci and reveals interactions with APOE genotypes. Arch Neurol 67(12), 1473–1484.

Khera, A.V., Chaffin, M., Aragam, K.G., Haas, M.E., Roselli, C., Choi, S.H., Natarajan, P., Lander, E.S., Lubitz, S.A., Ellinor, P.T., Kathiresan, S., 2018. Genome-wide polygenic scores for common diseases identify individuals with risk equivalent to monogenic mutations. Nat Genet 50(9), 1219–1224.

Koppel, J., Sousa, A., Gordon, M.L., Giliberto, L., Christen, E., Davies, P., 2018. Association Between Psychosis in Elderly Patients With Alzheimer Disease and Impaired Social Cognition. JAMA Psychiatry 75(6), 652–653.

Kunkle, B.W., Grenier-Boley, B., Sims, R., Bis, J.C., Damotte, V., Naj, A.C., Boland, A., Vronskaya, M., van der Lee, S.J., Amlie-Wolf, A., Bellenguez, C., Frizatti, A., Chouraki, V., Martin, E.R., Sleegers, K., Badarinarayan, N., Jakobsdottir, J., Hamilton-Nelson, K.L., Moreno-Grau, S., Olaso, R., Raybould, R., Chen, Y., Kuzma, A.B., Hiltunen, M., Morgan, T., Ahmad, S., Vardarajan, B.N., Epelbaum, J., Hoffmann, P., Boada, M., Beecham, G.W., Garnier, J.G., Harold, D., Fitzpatrick, A.L., Valladares, O., Moutet, M.L., Gerrish, A., Smith, A.V., Qu, L., Bacq, D., Denning, N., Jian, X., Zhao, Y., Del Zompo, M., Fox, N.C., Choi, S.H., Mateo, I., Hughes, J.T., Adams, H.H., Malamon, J., Sanchez-Garcia, F., Patel, Y., Brody, J.A., Dombroski, B.A., Naranjo, M.C.D., Daniilidou, M., Eiriksdottir, G., Mukherjee, S., Wallon, D., Uphill, J., Aspelund, T., Cantwell, L.B., Garzia, F., Galimberti, D., Hofer, E., Butkiewicz, M., Fin, B., Scarpini, E., Sarnowski, C., Bush, W.S., Meslage, S., Kornhuber, J., White, C.C., Song, Y., Barber, R.C., Engelborghs, S., Sordon, S., Voijnovic, D., Adams, P.M., Vandenberghe, R., Mayhaus, M., Cupples, L.A., Albert, M.S., De Deyn, P.P., Gu, W., Himali, J.J., Beekly, D., Squassina, A., Hartmann, A.M., Orellana, A., Blacker, D., Rodriguez-Rodriguez, E., Lovestone, S., Garcia, M.E., Doody, R.S., Munoz-Fernadez, C., Sussams, R., Lin, H., Fairchild, T.J., Benito, Y.A., Holmes, C., Karamujić-Čomić, H., Frosch, M.P., Thonberg, H., Maier, W., Roshchupkin, G., Ghetti, B., Giedraitis, V., Kawalia, A., Li, S., Huebinger, R.M., Kilander, L., Moebus, S., Hernández, I., Kamboh, M.I., Brundin, R., Turton, J., Yang, Q., Katz, M.J., Concari, L., Lord, J., Beiser, A.S., Keene, C.D., Helisalmi, S., Kloszewska, I., Kukull, W.A., Koivisto, A.M., Lynch, A., Tarraga, L., Larson, E.B., Haapasalo, A., Lawlor, B., Mosley, T.H., Lipton, R.B., Solfrizzi, V., Gill, M., Longstreth, W.T., Montine, T.J., Frisardi, V., Diez-Fairen, M., Rivadeneira, F., Petersen, R.C., Deramecourt, V., Alvarez, I., Salani, F., Ciaramella, A., Boerwinkle, E., Reiman, E.M., Fievet, N., Rotter, J.I., Reisch, J.S., Hanon, O., Cupidi, C., Andre Uitterlinden, A.G., Royall, D.R., Dufouil, C., Maletta, R.G., de Rojas, I., Sano, M., Brice, A., Cecchetti, R., George-Hyslop, P.S., Ritchie, K., Tsolaki, M., Tsuang, D.W., Dubois, B., Craig, D., Wu, C.K., Soininen, H., Avramidou, D., Albin, R.L., Fratiglioni, L., Germanou, A., Apostolova, L.G., Keller, L., Koutroumani, M., Arnold, S.E., Panza, F., Gkatzima, O., Asthana, S., Hannequin, D., Whitehead, P., Atwood, C.S., Caffarra, P., Hampel, H., Quintela, I., Carracedo, Á., Lannfelt, L., Rubinsztein, D.C., Barnes, L.L., Pasquier, F., Frölich, L., Barral, S., McGuinness, B., Beach, T.G., Johnston, J.A., Becker, J.T., Passmore, P., Bigio, E.H., Schott, J.M., Bird, T.D., Warren, J.D., Boeve, B.F., Lupton, M.K., Bowen, J.D., Proitsi, P., Boxer, A., Powell, J.F., Burke, J.R., Kauwe, J.S.K., Burns, J.M., Mancuso, M., Buxbaum, J.D., Bonuccelli, U., Cairns, N.J., McQuillin, A., Cao, C., Livingston, G., Carlson, C.S., Bass, N.J., Carlsson, C.M., Hardy, J., Carney, R.M., Bras, J., Carrasquillo, M.M., Guerreiro, R., Allen, M., Chui, H.C., Fisher, E., Masullo, C., Crocco, E.A., DeCarli, C., Bisceglio, G., Dick, M., Ma, L., Duara, R., Graff-Radford, N.R., Evans, D.A., Hodges, A., Faber, K.M., Scherer, M., Fallon, K.B., Riemenschneider, M., Fardo, D.W., Heun, R., Farlow, M.R., Kölsch, H., Ferris, S., Leber, M., Foroud, T.M., Heuser, I., Galasko, D.R., Giegling, I., Gearing, M., Hüll, M., Geschwind, D.H., Gilbert, J.R., Morris, J., Green, R.C., Mayo, K., Growdon, J.H., Feulner, T., Hamilton, R.L., Harrell, L.E., Drichel, D., Honig, L.S., Cushion, T.D., Huentelman, M.J., Hollingworth, P., Hulette, C.M., Hyman, B.T., Marshall, R., Jarvik, G.P., Meggy, A., Abner, E., Menzies, G.E., Jin, L.W., Leonenko, G., Real, L.M., Jun, G.R., Baldwin, C.T., Grozeva, D., Karydas, A., Russo, G., Kaye, J.A., Kim, R., Jessen, F., Kowall, N.W., Vellas, B., Kramer, J.H., Vardy, E., LaFerla, F.M., Jöckel, K.H., Lah, J.J., Dichgans, M., Leverenz, J.B., Mann, D., Levey, A.I., Pickering-Brown, S., Lieberman, A.P., Klopp, N., Lunetta, K.L., Wichmann, H.E., Lyketsos, C.G., Morgan, K., Marson, D.C., Brown, K., Martiniuk, F., Medway, C., Mash, D.C., Nöthen, M.M., Masliah, E., Hooper, N.M., McCormick, W.C., Daniele, A., McCurry, S.M., Bayer, A., McDavid, A.N., Gallacher, J., McKee, A.C., van den Bussche, H., Mesulam, M., Brayne, C., Miller, B.L., Riedel-Heller, S., Miller, C.A., Miller, J.W., Al-Chalabi, A., Morris, J.C., Shaw, C.E., Myers, A.J., Wiltfang, J., O’Bryant, S., Olichney, J.M., Alvarez, V., Parisi, J.E., Singleton, A.B., Paulson, H.L., Collinge, J., Perry, W.R., Mead, S., Peskind, E., Cribbs, D.H., Rossor, M., Pierce, A., Ryan, N.S., Poon, W.W., Nacmias, B., Potter, H., Sorbi, S., Quinn, J.F., Sacchinelli, E., Raj, A., Spalletta, G., Raskind, M., Caltagirone, C., Bossù, P., Orfei, M.D., Reisberg, B., Clarke, R., Reitz, C., Smith, A.D., Ringman, J.M., Warden, D., Roberson, E.D., Wilcock, G., Rogaeva, E., Bruni, A.C., Rosen, H.J., Gallo, M., Rosenberg, R.N., Ben-Shlomo, Y., Sager, M.A., Mecocci, P., Saykin, A.J., Pastor, P., Cuccaro, M.L., Vance, J.M., Schneider, J.A., Schneider, L.S., Slifer, S., Seeley, W.W., Smith, A.G., Sonnen, J.A., Spina, S., Stern, R.A., Swerdlow, R.H., Tang, M., Tanzi, R.E., Trojanowski, J.Q., Troncoso, J.C., Van Deerlin, V.M., Van Eldik, L.J., Vinters, H.V., Vonsattel, J.P., Weintraub, S., Welsh-Bohmer, K.A., Wilhelmsen, K.C., Williamson, J., Wingo, T.S., Woltjer, R.L., Wright, C.B., Yu, C.E., Yu, L., Saba, Y., Pilotto, A., Bullido, M.J., Peters, O., Crane, P.K., Bennett, D., Bosco, P., Coto, E., Boccardi, V., De Jager, P.L., Lleo, A., Warner, N., Lopez, O.L., Ingelsson, M., Deloukas, P., Cruchaga, C., Graff, C., Gwilliam, R., Fornage, M., Goate, A.M., Sanchez-Juan, P., Kehoe, P.G., Amin, N., Ertekin-Taner, N., Berr, C., Debette, S., Love, S., Launer, L.J., Younkin, S.G., Dartigues, J.F., Corcoran, C., Ikram, M.A., Dickson, D.W., Nicolas, G., Campion, D., Tschanz, J., Schmidt, H., Hakonarson, H., Clarimon, J., Munger, R., Schmidt, R., Farrer, L.A., Van Broeckhoven, C., C O’Donovan, M., DeStefano, A.L., Jones, L., Haines, J.L., Deleuze, J.F., Owen, M.J., Gudnason, V., Mayeux, R., Escott-Price, V., Psaty, B.M., Ramirez, A., Wang, L.S., Ruiz, A., van Duijn, C.M., Holmans, P.A., Seshadri, S., Williams, J., Amouyel, P., Schellenberg, G.D., Lambert, J.C., Pericak-Vance, M.A., Alzheimer Disease Genetics Consortium (ADGC), European Alzheimer’s Disease Initiative (EADI), Cohorts for Heart and Aging Research in Genomic Epidemiology Consortium (CHARGE), Genetic and Environmental Risk in AD/Defining Genetic, P.l.a.E.R.f.A.s.D.C.G.P., 2019a. Genetic meta-analysis of diagnosed Alzheimer’s disease identifies new risk loci and implicates Aβ, tau, immunity and lipid processing. Nat Genet 51(3), 414–430.

Kunkle, B.W., Grenier-Boley, B., Sims, R., Bis, J.C., Damotte, V., Naj, A.C., Boland, A., Vronskaya, M., van der Lee, S.J., Amlie-Wolf, A., Bellenguez, C., Frizatti, A., Chouraki, V., Martin, E.R., Sleegers, K., Badarinarayan, N., Jakobsdottir, J., Hamilton-Nelson, K.L., Moreno-Grau, S., Olaso, R., Raybould, R., Chen, Y., Kuzma, A.B., Hiltunen, M., Morgan, T., Ahmad, S., Vardarajan, B.N., Epelbaum, J., Hoffmann, P., Boada, M., Beecham, G.W., Garnier, J.G., Harold, D., Fitzpatrick, A.L., Valladares, O., Moutet, M.L., Gerrish, A., Smith, A.V., Qu, L., Bacq, D., Denning, N., Jian, X., Zhao, Y., Del Zompo, M., Fox, N.C., Choi, S.H., Mateo, I., Hughes, J.T., Adams, H.H., Malamon, J., Sanchez-Garcia, F., Patel, Y., Brody, J.A., Dombroski, B.A., Naranjo, M.C.D., Daniilidou, M., Eiriksdottir, G., Mukherjee, S., Wallon, D., Uphill, J., Aspelund, T., Cantwell, L.B., Garzia, F., Galimberti, D., Hofer, E., Butkiewicz, M., Fin, B., Scarpini, E., Sarnowski, C., Bush, W.S., Meslage, S., Kornhuber, J., White, C.C., Song, Y., Barber, R.C., Engelborghs, S., Sordon, S., Voijnovic, D., Adams, P.M., Vandenberghe, R., Mayhaus, M., Cupples, L.A., Albert, M.S., De Deyn, P.P., Gu, W., Himali, J.J., Beekly, D., Squassina, A., Hartmann, A.M., Orellana, A., Blacker, D., Rodriguez-Rodriguez, E., Lovestone, S., Garcia, M.E., Doody, R.S., Munoz-Fernadez, C., Sussams, R., Lin, H., Fairchild, T.J., Benito, Y.A., Holmes, C., Karamujić-Čomić, H., Frosch, M.P., Thonberg, H., Maier, W., Roshchupkin, G., Ghetti, B., Giedraitis, V., Kawalia, A., Li, S., Huebinger, R.M., Kilander, L., Moebus, S., Hernández, I., Kamboh, M.I., Brundin, R., Turton, J., Yang, Q., Katz, M.J., Concari, L., Lord, J., Beiser, A.S., Keene, C.D., Helisalmi, S., Kloszewska, I., Kukull, W.A., Koivisto, A.M., Lynch, A., Tarraga, L., Larson, E.B., Haapasalo, A., Lawlor, B., Mosley, T.H., Lipton, R.B., Solfrizzi, V., Gill, M., Longstreth, W.T., Montine, T.J., Frisardi, V., Diez-Fairen, M., Rivadeneira, F., Petersen, R.C., Deramecourt, V., Alvarez, I., Salani, F., Ciaramella, A., Boerwinkle, E., Reiman, E.M., Fievet, N., Rotter, J.I., Reisch, J.S., Hanon, O., Cupidi, C., Andre Uitterlinden, A.G., Royall, D.R., Dufouil, C., Maletta, R.G., de Rojas, I., Sano, M., Brice, A., Cecchetti, R., George-Hyslop, P.S., Ritchie, K., Tsolaki, M., Tsuang, D.W., Dubois, B., Craig, D., Wu, C.K., Soininen, H., Avramidou, D., Albin, R.L., Fratiglioni, L., Germanou, A., Apostolova, L.G., Keller, L., Koutroumani, M., Arnold, S.E., Panza, F., Gkatzima, O., Asthana, S., Hannequin, D., Whitehead, P., Atwood, C.S., Caffarra, P., Hampel, H., Quintela, I., Carracedo, Á., Lannfelt, L., Rubinsztein, D.C., Barnes, L.L., Pasquier, F., Frölich, L., Barral, S., McGuinness, B., Beach, T.G., Johnston, J.A., Becker, J.T., Passmore, P., Bigio, E.H., Schott, J.M., Bird, T.D., Warren, J.D., Boeve, B.F., Lupton, M.K., Bowen, J.D., Proitsi, P., Boxer, A., Powell, J.F., Burke, J.R., Kauwe, J.S.K., Burns, J.M., Mancuso, M., Buxbaum, J.D., Bonuccelli, U., Cairns, N.J., McQuillin, A., Cao, C., Livingston, G., Carlson, C.S., Bass, N.J., Carlsson, C.M., Hardy, J., Carney, R.M., Bras, J., Carrasquillo, M.M., Guerreiro, R., Allen, M., Chui, H.C., Fisher, E., Masullo, C., Crocco, E.A., DeCarli, C., Bisceglio, G., Dick, M., Ma, L., Duara, R., Graff-Radford, N.R., Evans, D.A., Hodges, A., Faber, K.M., Scherer, M., Fallon, K.B., Riemenschneider, M., Fardo, D.W., Heun, R., Farlow, M.R., Kölsch, H., Ferris, S., Leber, M., Foroud, T.M., Heuser, I., Galasko, D.R., Giegling, I., Gearing, M., Hüll, M., Geschwind, D.H., Gilbert, J.R., Morris, J., Green, R.C., Mayo, K., Growdon, J.H., Feulner, T., Hamilton, R.L., Harrell, L.E., Drichel, D., Honig, L.S., Cushion, T.D., Huentelman, M.J., Hollingworth, P., Hulette, C.M., Hyman, B.T., Marshall, R., Jarvik, G.P., Meggy, A., Abner, E., Menzies, G.E., Jin, L.W., Leonenko, G., Real, L.M., Jun, G.R., Baldwin, C.T., Grozeva, D., Karydas, A., Russo, G., Kaye, J.A., Kim, R., Jessen, F., Kowall, N.W., Vellas, B., Kramer, J.H., Vardy, E., LaFerla, F.M., Jöckel, K.H., Lah, J.J., Dichgans, M., Leverenz, J.B., Mann, D., Levey, A.I., Pickering-Brown, S., Lieberman, A.P., Klopp, N., Lunetta, K.L., Wichmann, H.E., Lyketsos, C.G., Morgan, K., Marson, D.C., Brown, K., Martiniuk, F., Medway, C., Mash, D.C., Nöthen, M.M., Masliah, E., Hooper, N.M., McCormick, W.C., Daniele, A., McCurry, S.M., Bayer, A., McDavid, A.N., Gallacher, J., McKee, A.C., van den Bussche, H., Mesulam, M., Brayne, C., Miller, B.L., Riedel-Heller, S., Miller, C.A., Miller, J.W., Al-Chalabi, A., Morris, J.C., Shaw, C.E., Myers, A.J., Wiltfang, J., O’Bryant, S., Olichney, J.M., Alvarez, V., Parisi, J.E., Singleton, A.B., Paulson, H.L., Collinge, J., Perry, W.R., Mead, S., Peskind, E., Cribbs, D.H., Rossor, M., Pierce, A., Ryan, N.S., Poon, W.W., Nacmias, B., Potter, H., Sorbi, S., Quinn, J.F., Sacchinelli, E., Raj, A., Spalletta, G., Raskind, M., Caltagirone, C., Bossù, P., Orfei, M.D., Reisberg, B., Clarke, R., Reitz, C., Smith, A.D., Ringman, J.M., Warden, D., Roberson, E.D., Wilcock, G., Rogaeva, E., Bruni, A.C., Rosen, H.J., Gallo, M., Rosenberg, R.N., Ben-Shlomo, Y., Sager, M.A., Mecocci, P., Saykin, A.J., Pastor, P., Cuccaro, M.L., Vance, J.M., Schneider, J.A., Schneider, L.S., Slifer, S., Seeley, W.W., Smith, A.G., Sonnen, J.A., Spina, S., Stern, R.A., Swerdlow, R.H., Tang, M., Tanzi, R.E., Trojanowski, J.Q., Troncoso, J.C., Van Deerlin, V.M., Van Eldik, L.J., Vinters, H.V., Vonsattel, J.P., Weintraub, S., Welsh-Bohmer, K.A., Wilhelmsen, K.C., Williamson, J., Wingo, T.S., Woltjer, R.L., Wright, C.B., Yu, C.E., Yu, L., Saba, Y., Pilotto, A., Bullido, M.J., Peters, O., Crane, P.K., Bennett, D., Bosco, P., Coto, E., Boccardi, V., De Jager, P.L., Lleo, A., Warner, N., Lopez, O.L., Ingelsson, M., Deloukas, P., Cruchaga, C., Graff, C., Gwilliam, R., Fornage, M., Goate, A.M., Sanchez-Juan, P., Kehoe, P.G., Amin, N., Ertekin-Taner, N., Berr, C., Debette, S., Love, S., Launer, L.J., Younkin, S.G., Dartigues, J.F., Corcoran, C., Ikram, M.A., Dickson, D.W., Nicolas, G., Campion, D., Tschanz, J., Schmidt, H., Hakonarson, H., Clarimon, J., Munger, R., Schmidt, R., Farrer, L.A., Van Broeckhoven, C., C O’Donovan, M., DeStefano, A.L., Jones, L., Haines, J.L., Deleuze, J.F., Owen, M.J., Gudnason, V., Mayeux, R., Escott-Price, V., Psaty, B.M., Ramirez, A., Wang, L.S., Ruiz, A., van Duijn, C.M., Holmans, P.A., Seshadri, S., Williams, J., Amouyel, P., Schellenberg, G.D., Lambert, J.C., Pericak-Vance, M.A., Consortium, A.D.G., Initiative, E.A.s.D., Heart, C.f., Consortium, A.R.i.G.E., Genetic, Environmental Risk in AD/Defining Genetic, P.l., Consortium, E.R.f.A.s.D., 2019b. Genetic meta-analysis of diagnosed Alzheimer’s disease identifies new risk loci and implicates Aβ, tau, immunity and lipid processing. Nat Genet 51(3), 414–430.

Lambert, J.C., Ibrahim-Verbaas, C.A., Harold, D., Naj, A.C., Sims, R., Bellenguez, C., DeStafano, A.L., Bis, J.C., Beecham, G.W., Grenier-Boley, B., Russo, G., Thorton-Wells, T.A., Jones, N., Smith, A.V., Chouraki, V., Thomas, C., Ikram, M.A., Zelenika, D., Vardarajan, B.N., Kamatani, Y., Lin, C.F., Gerrish, A., Schmidt, H., Kunkle, B., Dunstan, M.L., Ruiz, A., Bihoreau, M.T., Choi, S.H., Reitz, C., Pasquier, F., Cruchaga, C., Craig, D., Amin, N., Berr, C., Lopez, O.L., De Jager, P.L., Deramecourt, V., Johnston, J.A., Evans, D., Lovestone, S., Letenneur, L., Morón, F.J., Rubinsztein, D.C., Eiriksdottir, G., Sleegers, K., Goate, A.M., Fiévet, N., Huentelman, M.W., Gill, M., Brown, K., Kamboh, M.I., Keller, L., Barberger-Gateau, P., McGuiness, B., Larson, E.B., Green, R., Myers, A.J., Dufouil, C., Todd, S., Wallon, D., Love, S., Rogaeva, E., Gallacher, J., St George-Hyslop, P., Clarimon, J., Lleo, A., Bayer, A., Tsuang, D.W., Yu, L., Tsolaki, M., Bossù, P., Spalletta, G., Proitsi, P., Collinge, J., Sorbi, S., Sanchez-Garcia, F., Fox, N.C., Hardy, J., Deniz Naranjo, M.C., Bosco, P., Clarke, R., Brayne, C., Galimberti, D., Mancuso, M., Matthews, F., European Alzheimer’s Disease, I., Genetic, Environmental Risk in Alzheimer’s, D., Alzheimer’s Disease Genetic, C., Cohorts for, H., Aging Research in Genomic, E., Moebus, S., Mecocci, P., Del Zompo, M., Maier, W., Hampel, H., Pilotto, A., Bullido, M., Panza, F., Caffarra, P., Nacmias, B., Gilbert, J.R., Mayhaus, M., Lannefelt, L., Hakonarson, H., Pichler, S., Carrasquillo, M.M., Ingelsson, M., Beekly, D., Alvarez, V., Zou, F., Valladares, O., Younkin, S.G., Coto, E., Hamilton-Nelson, K.L., Gu, W., Razquin, C., Pastor, P., Mateo, I., Owen, M.J., Faber, K.M., Jonsson, P.V., Combarros, O., O’Donovan, M.C., Cantwell, L.B., Soininen, H., Blacker, D., Mead, S., Mosley, T.H., Jr., Bennett, D.A., Harris, T.B., Fratiglioni, L., Holmes, C., de Bruijn, R.F., Passmore, P., Montine, T.J., Bettens, K., Rotter, J.I., Brice, A., Morgan, K., Foroud, T.M., Kukull, W.A., Hannequin, D., Powell, J.F., Nalls, M.A., Ritchie, K., Lunetta, K.L., Kauwe, J.S., Boerwinkle, E., Riemenschneider, M., Boada, M., Hiltuenen, M., Martin, E.R., Schmidt, R., Rujescu, D., Wang, L.S., Dartigues, J.F., Mayeux, R., Tzourio, C., Hofman, A., Nöthen, M.M., Graff, C., Psaty, B.M., Jones, L., Haines, J.L., Holmans, P.A., Lathrop, M., Pericak-Vance, M.A., Launer, L.J., Farrer, L.A., van Duijn, C.M., Van Broeckhoven, C., Moskvina, V., Seshadri, S., Williams, J., Schellenberg, G.D., Amouyel, P., 2013a. Meta-analysis of 74,046 individuals identifies 11 new susceptibility loci for Alzheimer’s disease. Nat Genet 45(12), 1452–1458.

Lambert, J.C., Ibrahim-Verbaas, C.A., Harold, D., Naj, A.C., Sims, R., Bellenguez, C., DeStafano, A.L., Bis, J.C., Beecham, G.W., Grenier-Boley, B., Russo, G., Thorton-Wells, T.A., Jones, N., Smith, A.V., Chouraki, V., Thomas, C., Ikram, M.A., Zelenika, D., Vardarajan, B.N., Kamatani, Y., Lin, C.F., Gerrish, A., Schmidt, H., Kunkle, B., Dunstan, M.L., Ruiz, A., Bihoreau, M.T., Choi, S.H., Reitz, C., Pasquier, F., Cruchaga, C., Craig, D., Amin, N., Berr, C., Lopez, O.L., De Jager, P.L., Deramecourt, V., Johnston, J.A., Evans, D., Lovestone, S., Letenneur, L., Morón, F.J., Rubinsztein, D.C., Eiriksdottir, G., Sleegers, K., Goate, A.M., Fiévet, N., Huentelman, M.W., Gill, M., Brown, K., Kamboh, M.I., Keller, L., Barberger-Gateau, P., McGuiness, B., Larson, E.B., Green, R., Myers, A.J., Dufouil, C., Todd, S., Wallon, D., Love, S., Rogaeva, E., Gallacher, J., St George-Hyslop, P., Clarimon, J., Lleo, A., Bayer, A., Tsuang, D.W., Yu, L., Tsolaki, M., Bossù, P., Spalletta, G., Proitsi, P., Collinge, J., Sorbi, S., Sanchez-Garcia, F., Fox, N.C., Hardy, J., Deniz Naranjo, M.C., Bosco, P., Clarke, R., Brayne, C., Galimberti, D., Mancuso, M., Matthews, F., Moebus, S., Mecocci, P., Del Zompo, M., Maier, W., Hampel, H., Pilotto, A., Bullido, M., Panza, F., Caffarra, P., Nacmias, B., Gilbert, J.R., Mayhaus, M., Lannefelt, L., Hakonarson, H., Pichler, S., Carrasquillo, M.M., Ingelsson, M., Beekly, D., Alvarez, V., Zou, F., Valladares, O., Younkin, S.G., Coto, E., Hamilton-Nelson, K.L., Gu, W., Razquin, C., Pastor, P., Mateo, I., Owen, M.J., Faber, K.M., Jonsson, P.V., Combarros, O., O’Donovan, M.C., Cantwell, L.B., Soininen, H., Blacker, D., Mead, S., Mosley, T.H., Bennett, D.A., Harris, T.B., Fratiglioni, L., Holmes, C., de Bruijn, R.F., Passmore, P., Montine, T.J., Bettens, K., Rotter, J.I., Brice, A., Morgan, K., Foroud, T.M., Kukull, W.A., Hannequin, D., Powell, J.F., Nalls, M.A., Ritchie, K., Lunetta, K.L., Kauwe, J.S., Boerwinkle, E., Riemenschneider, M., Boada, M., Hiltuenen, M., Martin, E.R., Schmidt, R., Rujescu, D., Wang, L.S., Dartigues, J.F., Mayeux, R., Tzourio, C., Hofman, A., Nöthen, M.M., Graff, C., Psaty, B.M., Jones, L., Haines, J.L., Holmans, P.A., Lathrop, M., Pericak-Vance, M.A., Launer, L.J., Farrer, L.A., van Duijn, C.M., Van Broeckhoven, C., Moskvina, V., Seshadri, S., Williams, J., Schellenberg, G.D., Amouyel, P. E.A.s.D.I., Disease, G., Alzheimer’s, E.R.i., Consortium, A.s.D.G., Epidemiology, C.f.H., Genomic, A.R.i., 2013b. Meta-analysis of 74,046 individuals identifies 11 new susceptibility loci for Alzheimer’s disease. Nat Genet 45(12), 1452–1458.

Lambert, J.C., Ibrahim-Verbaas, C.A., Harold, D., Naj, A.C., Sims, R., Bellenguez, C., DeStafano, A.L., Bis, J.C., Beecham, G.W., Grenier-Boley, B., Russo, G., Thorton-Wells, T.A., Jones, N., Smith, A.V., Chouraki, V., Thomas, C., Ikram, M.A., Zelenika, D., Vardarajan, B.N., Kamatani, Y., Lin, C.F., Gerrish, A., Schmidt, H., Kunkle, B., Dunstan, M.L., Ruiz, A., Bihoreau, M.T., Choi, S.H., Reitz, C., Pasquier, F., Cruchaga, C., Craig, D., Amin, N., Berr, C., Lopez, O.L., De Jager, P.L., Deramecourt, V., Johnston, J.A., Evans, D., Lovestone, S., Letenneur, L., Moron, F.J., Rubinsztein, D.C., Eiriksdottir, G., Sleegers, K., Goate, A.M., Fievet, N., Huentelman, M.W., Gill, M., Brown, K., Kamboh, M.I., Keller, L., Barberger-Gateau, P., McGuiness, B., Larson, E.B., Green, R., Myers, A.J., Dufouil, C., Todd, S., Wallon, D., Love, S., Rogaeva, E., Gallacher, J., St George-Hyslop, P., Clarimon, J., Lleo, A., Bayer, A., Tsuang, D.W., Yu, L., Tsolaki, M., Bossù, P., Spalletta, G., Proitsi, P., Collinge, J., Sorbi, S., Sanchez-Garcia, F., Fox, N.C., Hardy, J., Deniz Naranjo, M.C., Bosco, P., Clarke, R., Brayne, C., Galimberti, D., Mancuso, M., Matthews, F., Moebus, S., Mecocci, P., Del Zompo, M., Maier, W., Hampel, H., Pilotto, A., Bullido, M., Panza, F., Caffarra, P., Nacmias, B., Gilbert, J.R., Mayhaus, M., Lannefelt, L., Hakonarson, H., Pichler, S., Carrasquillo, M.M., Ingelsson, M., Beekly, D., Alvarez, V., Zou, F., Valladares, O., Younkin, S.G., Coto, E., Hamilton-Nelson, K.L., Gu, W., Razquin, C., Pastor, P., Mateo, I., Owen, M.J., Faber, K.M., Jonsson, P.V., Combarros, O., O’Donovan, M.C., Cantwell, L.B., Soininen, H., Blacker, D., Mead, S., Mosley, T.H., Bennett, D.A., Harris, T.B., Fratiglioni, L., Holmes, C., de Bruijn, R.F., Passmore, P., Montine, T.J., Bettens, K., Rotter, J.I., Brice, A., Morgan, K., Foroud, T.M., Kukull, W.A., Hannequin, D., Powell, J.F., Nalls, M.A., Ritchie, K., Lunetta, K.L., Kauwe, J.S., Boerwinkle, E., Riemenschneider, M., Boada, M., Hiltuenen, M., Martin, E.R., Schmidt, R., Rujescu, D., Wang, L.S., Dartigues, J.F., Mayeux, R., Tzourio, C., Hofman, A., Nöthen, M.M., Graff, C., Psaty, B.M., Jones, L., Haines, J.L., Holmans, P.A., Lathrop, M., Pericak-Vance, M.A., Launer, L.J., Farrer, L.A., van Duijn, C.M., Van Broeckhoven, C., Moskvina, V., Seshadri, S., Williams, J., Schellenberg, G.D., Amouyel, P., (EADI), E.A.s.D.I., Disease, G.a.E.R.i.A.s., Consortium, A.s.D.G., Epidemiology, C.f.H.a.A.R.i.G., 2013c. Meta-analysis of 74,046 individuals identifies 11 new susceptibility loci for Alzheimer’s disease. Nat Genet 45(12), 1452–1458.

Leonenko, G., Sims, R., Shoai, M., Frizzati, A., Bossù, P., Spalletta, G., Fox, N.C., Williams, J., Hardy, J., Escott-Price, V., consortium, G., 2019. Polygenic risk and hazard scores for Alzheimer’s disease prediction. Ann Clin Transl Neurol 6(3), 456–465.

Marioni, R.E., Harris, S.E., Zhang, Q., McRae, A.F., Hagenaars, S.P., Hill, W.D., Davies, G., Ritchie, C.W., Gale, C.R., Starr, J.M., Goate, A.M., Porteous, D.J., Yang, J., Evans, K.L., Deary, I.J., Wray, N.R., Visscher, P.M., 2018. GWAS on family history of Alzheimer’s disease. Transl Psychiatry 8(1), 99.

Martin, A.R., Kanai, M., Kamatani, Y., Okada, Y., Neale, B.M., Daly, M.J., 2019. Clinical use of current polygenic risk scores may exacerbate health disparities. Nat Genet 51(4), 584–591.

Mayeux, R., Stern, Y., 2012. Epidemiology of Alzheimer disease. Cold Spring Harb Perspect Med 2(8).

Naj, A.C., Jun, G., Beecham, G.W., Wang, L.S., Vardarajan, B.N., Buros, J., Gallins, P.J., Buxbaum, J.D., Jarvik, G.P., Crane, P.K., Larson, E.B., Bird, T.D., Boeve, B.F., Graff-Radford, N.R., De Jager, P.L., Evans, D., Schneider, J.A., Carrasquillo, M.M., Ertekin-Taner, N., Younkin, S.G., Cruchaga, C., Kauwe, J.S., Nowotny, P., Kramer, P., Hardy, J., Huentelman, M.J., Myers, A.J., Barmada, M.M., Demirci, F.Y., Baldwin, C.T., Green, R.C., Rogaeva, E., St George-Hyslop, P., Arnold, S.E., Barber, R., Beach, T., Bigio, E.H., Bowen, J.D., Boxer, A., Burke, J.R., Cairns, N.J., Carlson, C.S., Carney, R.M., Carroll, S.L., Chui, H.C., Clark, D.G., Corneveaux, J., Cotman, C.W., Cummings, J.L., DeCarli, C., DeKosky, S.T., Diaz-Arrastia, R., Dick, M., Dickson, D.W., Ellis, W.G., Faber, K.M., Fallon, K.B., Farlow, M.R., Ferris, S., Frosch, M.P., Galasko, D.R., Ganguli, M., Gearing, M., Geschwind, D.H., Ghetti, B., Gilbert, J.R., Gilman, S., Giordani, B., Glass, J.D., Growdon, J.H., Hamilton, R.L., Harrell, L.E., Head, E., Honig, L.S., Hulette, C.M., Hyman, B.T., Jicha, G.A., Jin, L.W., Johnson, N., Karlawish, J., Karydas, A., Kaye, J.A., Kim, R., Koo, E.H., Kowall, N.W., Lah, J.J., Levey, A.I., Lieberman, A.P., Lopez, O.L., Mack, W.J., Marson, D.C., Martiniuk, F., Mash, D.C., Masliah, E., McCormick, W.C., McCurry, S.M., McDavid, A.N., McKee, A.C., Mesulam, M., Miller, B.L., Miller, C.A., Miller, J.W., Parisi, J.E., Perl, D.P., Peskind, E., Petersen, R.C., Poon, W.W., Quinn, J.F., Rajbhandary, R.A., Raskind, M., Reisberg, B., Ringman, J.M., Roberson, E.D., Rosenberg, R.N., Sano, M., Schneider, L.S., Seeley, W., Shelanski, M.L., Slifer, M.A., Smith, C.D., Sonnen, J.A., Spina, S., Stern, R.A., Tanzi, R.E., Trojanowski, J.Q., Troncoso, J.C., Van Deerlin, V.M., Vinters, H.V., Vonsattel, J.P., Weintraub, S., Welsh-Bohmer, K.A., Williamson, J., Woltjer, R.L., Cantwell, L.B., Dombroski, B.A., Beekly, D., Lunetta, K.L., Martin, E.R., Kamboh, M.I., Saykin, A.J., Reiman, E.M., Bennett, D.A., Morris, J.C., Montine, T.J., Goate, A.M., Blacker, D., Tsuang, D.W., Hakonarson, H., Kukull, W.A., Foroud, T.M., Haines, J.L., Mayeux, R., Pericak-Vance, M.A., Farrer, L.A., Schellenberg, G.D., 2011. Common variants at MS4A4/MS4A6E, CD2AP, CD33 and EPHA1 are associated with late-onset Alzheimer’s disease. Nat Genet 43(5), 436–441.

Naj, A.C., Jun, G., Reitz, C., Kunkle, B.W., Perry, W., Park, Y.S., Beecham, G.W., Rajbhandary, R.A., Hamilton-Nelson, K.L., Wang, L.S., Kauwe, J.S., Huentelman, M.J., Myers, A.J., Bird, T.D., Boeve, B.F., Baldwin, C.T., Jarvik, G.P., Crane, P.K., Rogaeva, E., Barmada, M.M., Demirci, F.Y., Cruchaga, C., Kramer, P.L., Ertekin-Taner, N., Hardy, J., Graff-Radford, N.R., Green, R.C., Larson, E.B., St George-Hyslop, P.H., Buxbaum, J.D., Evans, D.A., Schneider, J.A., Lunetta, K.L., Kamboh, M.I., Saykin, A.J., Reiman, E.M., De Jager, P.L., Bennett, D.A., Morris, J.C., Montine, T.J., Goate, A.M., Blacker, D., Tsuang, D.W., Hakonarson, H., Kukull, W.A., Foroud, T.M., Martin, E.R., Haines, J.L., Mayeux, R.P., Farrer, L.A., Schellenberg, G.D., Pericak-Vance, M.A., Albert, M.S., Albin, R.L., Apostolova, L.G., Arnold, S.E., Barber, R., Barnes, L.L., Beach, T.G., Becker, J.T., Beekly, D., Bigio, E.H., Bowen, J.D., Boxer, A., Burke, J.R., Cairns, N.J., Cantwell, L.B., Cao, C., Carlson, C.S., Carney, R.M., Carrasquillo, M.M., Carroll, S.L., Chui, H.C., Clark, D.G., Corneveaux, J., Cribbs, D.H., Crocco, E.A., DeCarli, C., DeKosky, S.T., Dick, M., Dickson, D.W., Duara, R., Faber, K.M., Fallon, K.B., Farlow, M.R., Ferris, S., Frosch, M.P., Galasko, D.R., Ganguli, M., Gearing, M., Geschwind, D.H., Ghetti, B., Gilbert, J.R., Glass, J.D., Growdon, J.H., Hamilton, R.L., Harrell, L.E., Head, E., Honig, L.S., Hulette, C.M., Hyman, B.T., Jicha, G.A., Jin, L.W., Karydas, A., Kaye, J.A., Kim, R., Koo, E.H., Kowall, N.W., Kramer, J.H., LaFerla, F.M., Lah, J.J., Leverenz, J.B., Levey, A.I., Li, G., Lieberman, A.P., Lin, C.F., Lopez, O.L., Lyketsos, C.G., Mack, W.J., Martiniuk, F., Mash, D.C., Masliah, E., McCormick, W.C., McCurry, S.M., McDavid, A.N., McKee, A.C., Mesulam, M., Miller, B.L., Miller, C.A., Miller, J.W., Murrell, J.R., Olichney, J.M., Pankratz, V.S., Parisi, J.E., Paulson, H.L., Peskind, E., Petersen, R.C., Pierce, A., Poon, W.W., Potter, H., Quinn, J.F., Raj, A., Raskind, M., Reisberg, B., Ringman, J.M., Roberson, E.D., Rosen, H.J., Rosenberg, R.N., Sano, M., Schneider, L.S., Seeley, W.W., Smith, A.G., Sonnen, J.A., Spina, S., Stern, R.A., Tanzi, R.E., Thornton-Wells, T.A., Trojanowski, J.Q., Troncoso, J.C., Valladares, O., Van Deerlin, V.M., Van Eldik, L.J., Vardarajan, B.N., Vinters, H.V., Vonsattel, J.P., Weintraub, S., Welsh-Bohmer, K.A., Williamson, J., Wishnek, S., Woltjer, R.L., Wright, C.B., Younkin, S.G., Yu, C.E., Yu, L., Consortium, A.D.G., 2014. Effects of multiple genetic loci on age at onset in late-onset Alzheimer disease: a genome-wide association study. JAMA Neurol 71(11), 1394–1404.

Olarte, L., Schupf, N., Lee, J.H., Tang, M.X., Santana, V., Williamson, J., Maramreddy, P., Tycko, B., Mayeux, R., 2006. Apolipoprotein E epsilon4 and age at onset of sporadic and familial Alzheimer disease in Caribbean Hispanics. Arch Neurol 63(11), 1586–1590.

Purcell, S.M., Wray, N.R., Stone, J.L., Visscher, P.M., O’Donovan, M.C., Sullivan, P.F., Sklar, P., Consortium, I.S., 2009. Common polygenic variation contributes to risk of schizophrenia and bipolar disorder. Nature 460(7256), 748–752.

R Core Team, 2019. R: A language and environment for statistical computing.

Rajabli, F., Feliciano, B.E., Celis, K., Hamilton-Nelson, K.L., Whitehead, P.L., Adams, L.D., Bussies, P.L., Manrique, C.P., Rodriguez, A., Rodriguez, V., Starks, T., Byfield, G.E., Sierra Lopez, C.B., McCauley, J.L., Acosta, H., Chinea, A., Kunkle, B.W., Reitz, C., Farrer, L.A., Schellenberg, G.D., Vardarajan, B.N., Vance, J.M., Cuccaro, M.L., Martin, E.R., Haines, J.L., Byrd, G.S., Beecham, G.W., Pericak-Vance, M.A., 2018. Ancestral origin of ApoE epsilon4 Alzheimer disease risk in Puerto Rican and African American populations. PLoS Genet 14(12), e1007791.

Rasmussen, K.L., Tybjaerg-Hansen, A., Nordestgaard, B.G., Frikke-Schmidt, R., 2018. Absolute 10-year risk of dementia by age, sex and APOE genotype: a population-based cohort study. CMAJ 190(35), E1033-E1041.

Sebastiani, P., Gurinovich, A., Nygaard, M., Sasaki, T., Sweigart, B., Bae, H., Andersen, S.L., Villa, F., Atzmon, G., Christensen, K., Arai, Y., Barzilai, N., Puca, A., Christiansen, L., Hirose, N., Perls, T.T., 2019. APOE Alleles and Extreme Human Longevity. J Gerontol A Biol Sci Med Sci 74(1), 44–51.

Tesi, N., van der Lee, S.J., Hulsman, M., Jansen, I.E., Stringa, N., van Schoor, N., Meijers-Heijboer, H., Huisman, M., Scheltens, P., Reinders, M.J.T., van der Flier, W.M., Holstege, H., 2019. Centenarian controls increase variant effect sizes by an average twofold in an extreme case-extreme control analysis of Alzheimer’s disease. Eur J Hum Genet 27(2), 244–253.

